# Clinicopathological and Imaging Features of Primary Biliary Cholangitis

**DOI:** 10.1101/2022.04.29.22274511

**Authors:** Mengdan Xie, Custon Tafadzwa Nyabanga, Mantej Sehmbhi, Nour Al Khalili, Christy Chon, Amita Kamath, Amreen M. Dinani, Ilan S. Weisberg

**Affiliations:** Department of Medicine, Mount Sinai Morningside and West Hospitals; Division of Gastroenterology and Hepatology, Department of Medicine, Mount Sinai Beth Israel, Morningside and West Hospitals; Division of liver Diseases, Icahn School of Medicine at Mount Sinai; Department of Radiology, Mount Sinai Hospital & Mount Sinai Beth Israel, Morningside and West Hospitals

**Keywords:** Primary biliary cholangitis, image, periportal halo sign, gender, antimitochondrial antibody

## Abstract

**Background and Aims:** Primary biliary cholangitis (PBC) is a chronic inflammatory autoimmune disease of the biliary epithelial cells, causing slow progression of cholestasis and fibrosis. The aim of the study is to summarize the imaging features of PBC and examine the correlation between its clinicopathological and radiologic features, elucidating the specific clinicopathological and radiologic differences between male and female PBC patients, as well as between AMA-positive and AMA-negative PBC patients.

**Methods:** Demographic, laboratory, radiologic and survival data were collected retrospectively for patients diagnosed and treated for PBC at the Mount Sinai Health System between 2016 and 2020 with at least one ultrasound, CT, or MRI of abdomen available for assessment. Biochemical and radiologic data were compared between male v.s. female groups, and AMA-positive v.s. AMA-negative group.

**Results:** A total of 273 patients diagnosed with PBC were included. Non-specific hepatic parenchymal disease, cirrhosis, cholelithiasis, splenomegaly, lymphadenopathy, benign liver masses/cysts were the most common features on abdominal images. 24% of PBC patients had biliary tree abnormalities on MRI. LAD was reported in 38-47% of PBC patients. There was no significant difference in age, race, or ethnicity between the sexes of patients with PBC, however imaging characteristics of portal hypertension were more frequently seen in men. AMA-negative PBC patients had similar distributions of age, race/ethnicity and survival to AMA-positive PBC patients.

**Conclusion:** Our study summarized the clinicopathological and imaging features of PBC and distinct subgroups. This would assist in the diagnosis of PBC and avoid over-testing in real-world practice.

## Introduction

Primary biliary cholangitis (PBC) is a chronic autoimmune disease characterized by progressive small bile duct destruction, cholestasis and variable progression to cirrhosis. It is most commonly seen in women, with a female-to-male prevalence ratio thought to be 9:1^1^, but with recent data suggesting regional variation in this balance with ratios varying between 2:1 and 4:1^2^. Though the pathogenesis of PBC is understood to involve T-cell mediated biliary injury in all patients, it is a heterogeneous disease, with clinical manifestations varying from chronically asymptomatic disease to cirrhosis within years.

The role of imaging in PBC has largely been limited to the exclusion of other disorders and monitoring of hepatic fibrosis^3^. Clinical guidelines from both the American Association for the Study of Liver Diseases (AASLD) and the European Association for the Study of the Liver (EASL) recommend that imaging be performed as part of the diagnostic evaluation of cholestasis, but with the aim of excluding extrahepatic causes of cholestasis rather than seeking direct radiological evidence of PBC^4,5^.

Ultrasound imaging is central also to monitoring for progression of fibrosis and surveillance of hepatocellular carcinoma, but imaging findings in the later stages of PBC resemble fibrotic changes from viral and alcoholic liver diseases^3^, limiting the utility of these studies in the specific diagnosis of PBC. Magnetic resonance (MR) examinations have shown some promise in identifying specific features of PBC, such as the periportal halo sign, but these are typically seen late in the disease^6^. Abdominal lymphadenopathy^7-9^ is seen in PBC but is non-specific^6^. Studies of computed tomography (CT) images in PBC have long shown a tendency for earlier development of sequelae of portal hypertension than would be expected for the macroscopic degree of hepatic fibrosis^9,10^, although they are insufficiently specific to make a definitive diagnosis. However, it has been suggested that some of these features are useful in predicting advanced fibrosis^11^, and thus may obviate the need for biopsy.

One drawback of the existing literature on imaging features of PBC is the relative paucity of clinical information and clinicopathological correlation in these studies. While most of the literature discusses the role of imaging in PBC in relation to monitoring disease progression and the early detection of complications, very few studies examine the radiographic differences that may exist between distinct subgroups within the overall PBC population, such as differences between demographic groups or between those with atypical biochemical profiles. Additionally, many of the existing studies were led by radiologists, which could introduce the potential for observer bias, limiting the scope of reported radiological findings within PBC in real-world clinical practice.

As such, the overall aim of our study was to summarize the imaging features of PBC and examine the correlation between its clinicopathological and radiologic features. We also aimed to elucidate the specific demographic, biochemical, radiologic, and survival differences between male and female PBC patients, as well as between AMA-positive and AMA-negative PBC patients.

## Materials and methods

### Patient population

A retrospective review of electronic medical records at the Mount Sinai Health System from January 2016 through December 2020 revealed 273 patients with a diagnosis of PBC and at least one ultrasound (US), CT, or MRI of the abdomen available for assessment. We confirmed that all included patients met the criteria for a diagnosis of PBC as per AASLD guidance, which requires the presence of at least two of these three criteria: 1) positive antimitochondrial antibody; 2) elevated alkaline phosphatase; 3) liver biopsy confirming diagnosis^5^. Patients with coexistent viral liver disease, primary sclerosing cholangitis, liver metastases, or other diffuse liver diseases were excluded. Demographic, histologic, laboratory and radiologic data were collected for analysis. This study was performed with the approval of the Institutional Review Board (IRB) of the Icahn School of Medicine at Mount Sinai. Written consent was waived as per IRB protocol.

### Histological evaluation

114 patients had liver biopsies performed within the Mount Sinai Health System. Liver histology had been evaluated by specialized hepatic pathologists from the Mount Sinai Department of Pathology. Histopathologic proof of PBC was based on a chronic, nonsuppurative cholangitis mainly affecting the interlobular and septal bile ducts. Histologic lesions had been divided into 4 stages depending on the severity and extent of inflammation and fibrosis: stage 1, portal inflammation with or without florid bile duct lesions; stage 2, interface hepatitis with periportal lesions extending into the hepatic parenchyma; stage 3, bridging inflammation with formation of fibrous septa; and stage 4, cirrhosis with the existence of regenerative nodules^5^.

### Imaging technique and analysis

Transabdominal grayscale ultrasound examinations of the abdomen and retroperitoneum had been performed by sonogram technicians as per standard protocol. Helical multiphase CT scans of the abdomen had been performed from the lung bases to the pubic symphysis with 100 mL of Isovue-300 intravenous contrast. Nonenhanced and contrast-enhanced images of the abdomen were obtained in all cases. MRI of the abdomen or magnetic resonance cholangiopancreatography (MRCP) had been performed with the following sequences: axial and coronal T2 half-fourier acquisition single-shot turbo spin echo imaging (HASTE), axial T2 fat saturated, axial diffusion, axial T1 in and out of phase, thick slab and 3-dimensional (3D) MRCP, axial and coronal 3D T1 weighted before and after 10 mL intravenous Eovist administration. MR elastography had also been performed in some cases. Radiographic evidence of portal hypertension included: gastric/esophageal varices, recanalized paraumbilical vein, splenorenal shunt, splenomegaly or ascites.

The review of PBC images were performed by one or two abdominal radiologists under real-world clinical practice. We extracted the following features retrospectively from imaging reports: liver cirrhosis, hepatomegaly, hepatic masses, hepatic steatosis, periportal halo sign (for MR imaging), cholelithiasis, gallbladder sludge, gallbladder wall abnormalities, biliary tree abnormalities, ascites, splenomegaly, portosystemic collaterals, lymphadenopathy (defined as ≥1 cm in the short axis). Other intra-abdominal abnormalities were recorded separately. In patients with more than one imaging report, only the most recent report for each of the modalities (US or CT or MRI) was included for final analysis.

### Statistical analyses

All statistical analyses and data visualization were performed using RStudio version 3.6.1 and STATA 15. Continuous variables were presented as mean ± standard deviation (SD). Proportions (as percentages) were given for categorical or ordinal data. Normality was evaluated with the Kolmogorov-Smirnov test. The *t*-test was utilized to test for a difference in means between groups for continuous variables, while Chi-squared or Fisher’s exact tests were used to compare categorical data between groups.

## Results

### Clinicopathological features

A total of 273 patients diagnosed with PBC were included (table 1). The study population included 243 (89%) female and 41 (15%) AMA-negative PBC patients, with a mean age of 64.5+/-1.5 years. Liver biopsy was performed in 114 patients: 23 (20.2%) showed stage 1 PBC, 18 (15.8%) stage 2, 43 (37.8%) stage 3 and 11 (9.7%) stage 4; 19 patients had no evidence of PBC on biopsy (PBC was confirmed by elevated ALP, positive AMA, and exclusion of other liver diseases). Fifty-four patients had evidence of autoimmune hepatitis overlap syndrome. Two hundred and thirty PBC patients had detailed radiographic results for analysis: 172 (75%) had US, 95 (41%) had CT, and 129 (56%) had MRI results.

**Table 1.** Clinicopathologic Features of PBC patients

|  |  | ALL | WITH IMAGE | US | CT | MRI |
| --- | --- | --- | --- | --- | --- | --- |
| **N** |  | 273 | 230 | 172 | 95 | 129 |
| **AGE (YO, 95%CI)** |  | 64.5<br>(63.0-66.0) | 64.3<br>(62.7-66.0) | 64.3<br>(62.3-66.2) | 68.6<br>(66.2-70.9) | 63.0<br>(61.0-65.0) |
| **GENDER, N (%)** | Male | 30(11.0) | 27 (11.7) | 25 (14.5) | 15 (15.8) | 16 (12.4) |
|  | Female | 243(89.1) | 203 (88.3) | 147 (85.5) | 80 (84.2) | 113 (87.6) |
| **RACE/ETHNICITY, N (%)** | African American | 20(7.3) | 16 (7) | 12 (7) | 8 (8.4) | 8 (6.2) |
|  | White | 126(46.2) | 106 (46.1) | 80 (46.5) | 47 (49.5) | 62 (48.1) |
|  | Asian | 12(4.4) | 12 (5.2) | 7 (4.1) | 6 (6.3) | 4 (3.1) |
|  | Other | 78(28.6) | 66 (28.7) | 54 (31.4) | 32 (33.7) | 40 (31) |
|  | Unknown | 37(13.6) | 30 (13) | 19 (11.1) | 2 (2.1) | 15 (11.6) |
| **ALP, N (%)** | Elevated | 253(92.7) | 225 (97.8) | 167 (97.1) | 94 (99) | 127 (98.5) |
|  | Non-elevated | 19(7.0) | 5 (2.2) | 5 (2.9) | 1 (1.1) | 2 (1.6) |
| **AMA, N (%)** | Positive | 230(84.3) | 207 (90) | 154 (89.5) | 86 (90.5) | 113 (87.6) |
|  | Negative | 41(15.0) | 22 (9.6) | 18 (10.5) | 9 (9.5) | 15 (11.6) |
|  | N/A | 2(0.7) | 1 (0.4) | 0 (0) | 0 (0) | 1 (0.8) |
| **LIVER BIOPSY, N (%)** | Done | 114(41.8) | 104 (45.2) | 81 (47.1) | 43 (45.3) | 62 (48.1) |
|  | N/A | 159(58.2) | 126 (54.8) | 91 (52.9) | 52 (54.7) | 67 (51.9) |
| **STAGE OF PBC ON BIOPSY, N (%)** | 1 | 23(20.1) | 22 (21.2) | 17 (21) | 6 (14) | 11 (17.7) |
|  | 2 | 18(15.8) | 16 (15.4) | 14 (17.3) | 9 (20.9) | 10 (16.1) |
|  | 3 | 43(37.8) | 42 (40.4) | 32 (39.5) | 17 (39.5) | 26 (41.9) |
|  | 4 | 11(9.7) | 11 (10.6) | 9 (11.1) | 5 (11.6) | 6 (9.7) |
|  | Not PBC | 19(16.7) | 13 (12.5) | 9 (11.1) | 6 (14) | 9 (14.5) |
| **OVERLAP WITH AIH, N (%)** | No | 219(80.2) | 187 (81.3) | 135 (78.5) | 76 (80) | 107 (83) |
|  | Yes | 54(19.8) | 43 (18.7) | 37 (21.5) | 19 (20) | 22 (17.1) |

### Imaging features

US, CT, MRI features are summarized and compared in table 2 and figure. Non-specific hepatic parenchymal disease (51%), cirrhosis (32%), cholelithiasis (20%), splenomegaly (12%) and lymphadenopathy (LAD) (11%) were the most common features found on US in patients with PBC. Cirrhosis (50%), LAD (38%), splenomegaly (28%), benign liver masses/cysts (18%) and cholelithiasis (16%) were the most common features on CT in patients with PBC. Imaging evidence of liver cirrhosis and portal hypertension were more frequently observed in the CT group (49.5% and 48.4% respectively), which may suggest that this population had relatively more advanced PBC. LAD (47%), cirrhosis (41%), benign liver masses/cysts (36%), splenomegaly (23%) and cholelithiasis (11%) were the most common features on MRI in patients with PBC. 24% of PBC patients had biliary tree abnormalities on MRI, including intrahepatic (7%) and extrahepatic (2%) biliary duct dilatation, common bile duct dilatation (8%), intrahepatic duct stricture (2%), or other biliary tree irregularities (6%). LAD was reported in 38-47% of PBC patients, commonly involving periportal, porta hepatis, portacaval and peripancreatic lymph nodes. Notably, 147 (85.5%) ultrasound reports did not mention lymphadenopathy in our sample. No imaging evidence of hepatocellular carcinoma (HCC) was found in our cohort.

**Table 2.**
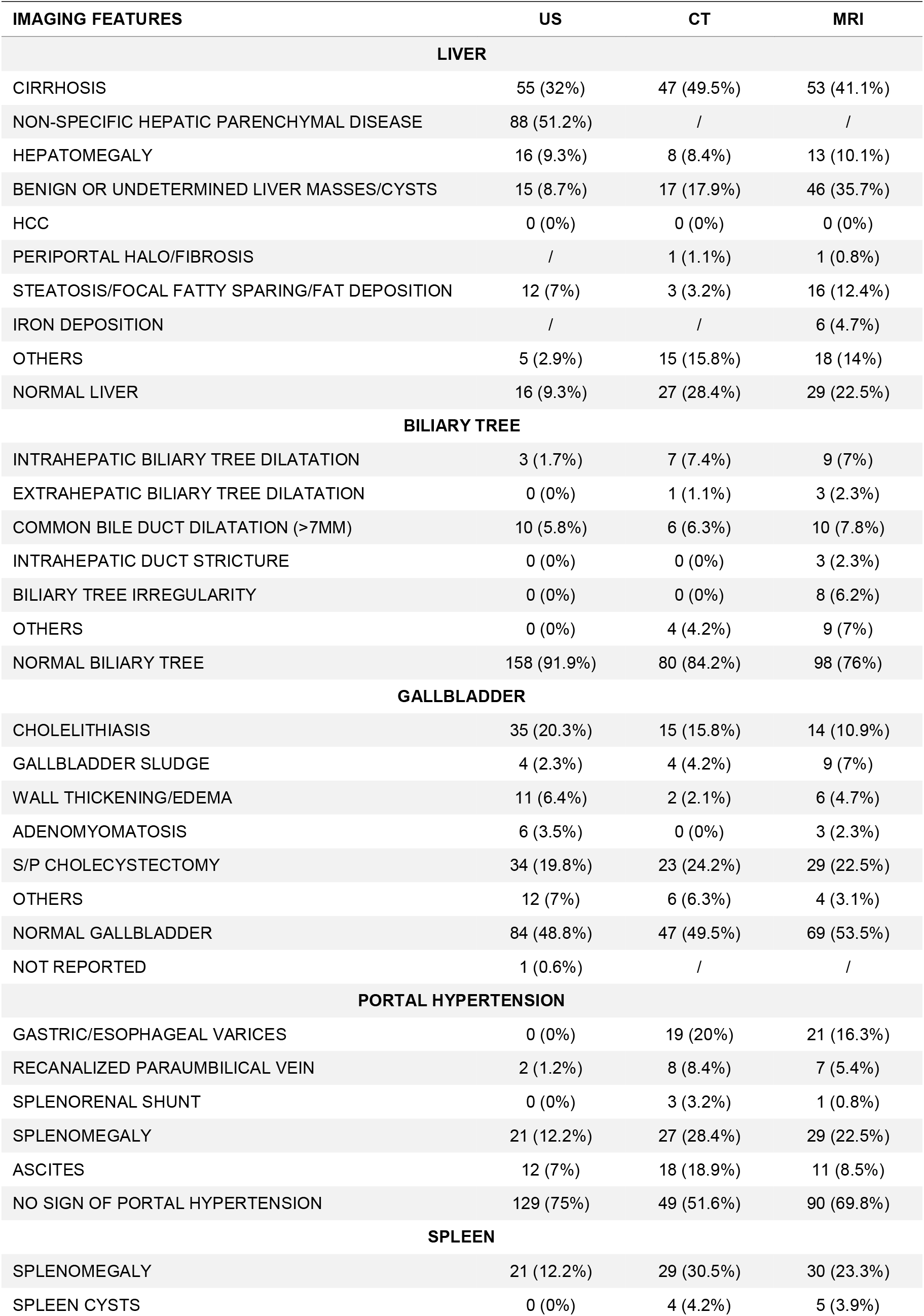
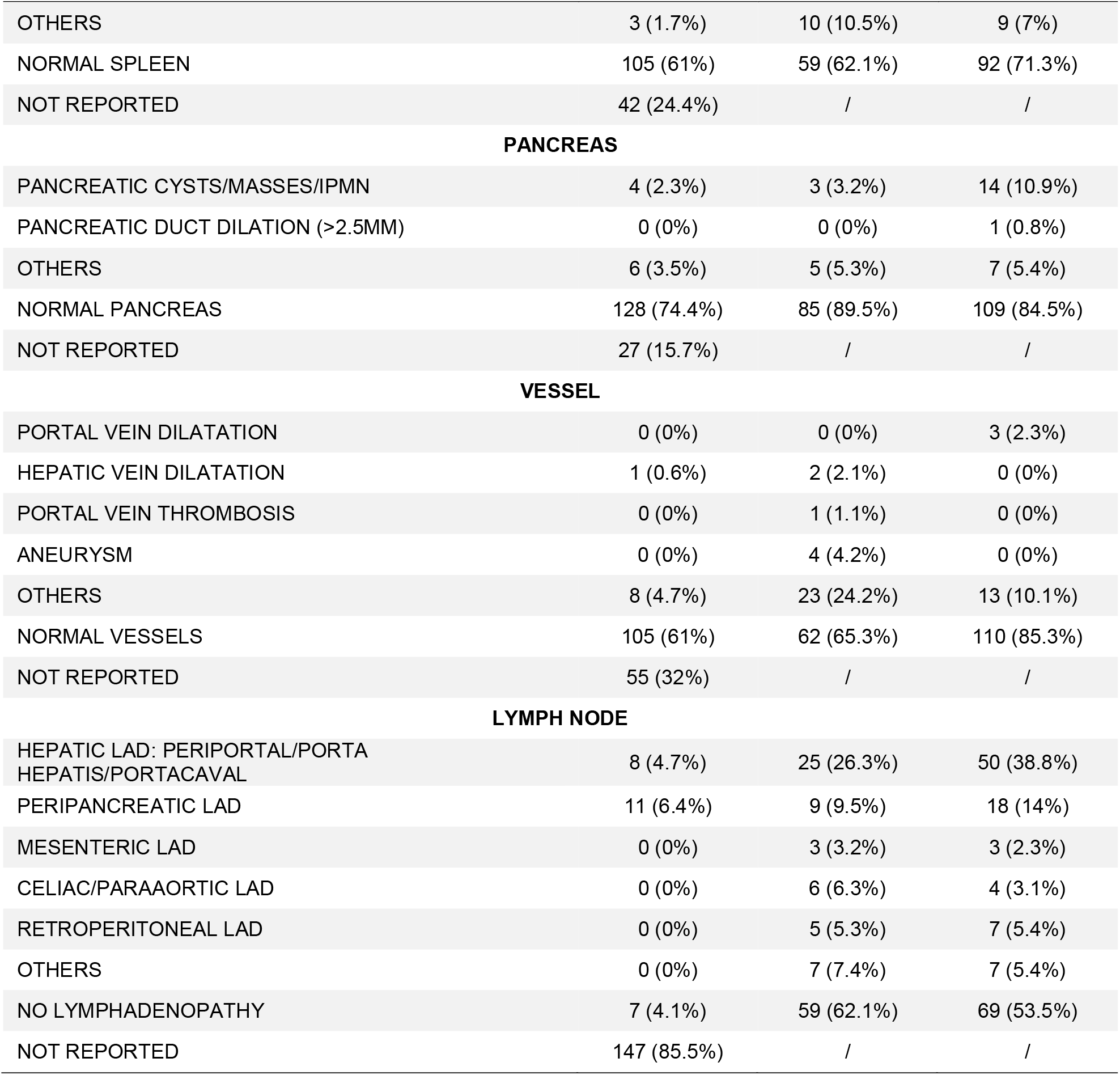
Imaging features of PBC patients

### Comparison between men and women with PBC

There was no significant difference in age, race, or ethnicity between the sexes of patients with PBC (table 3). Alkaline phosphatase (ALP) was elevated in 97% of men vs. 92% of women. The mean level of ALP was higher in men than in women (442 vs. 316 u/L, *p* = 0.058). Antimitochondrial antibody (AMA) was positive in 73% of men vs. 69% of women. Liver biopsies were performed in 11/30 (37%) males and 103/243 (42%) females. Early PBC (stage 1 or 2) was seen in 12.5% of men vs. 46% of women while advanced PBC (stage 3 or 4) was seen in 88% of men vs. 54% of women (*p* = 0.068). Of note, 23.3% of men vs. 19.3% of women were diagnosed with autoimmune hepatitis overlap syndrome with no statistically significant difference. 231 patients had radiographic studies for assessment. Radiographic evidence for cirrhosis was similar in men and women (48% vs. 42%); however, imaging characteristics of portal hypertension (splenomegaly, ascites, and varices) were more frequently seen in men (55.5% vs. 35.9%, *p* < 0.05).

**Table 3.**
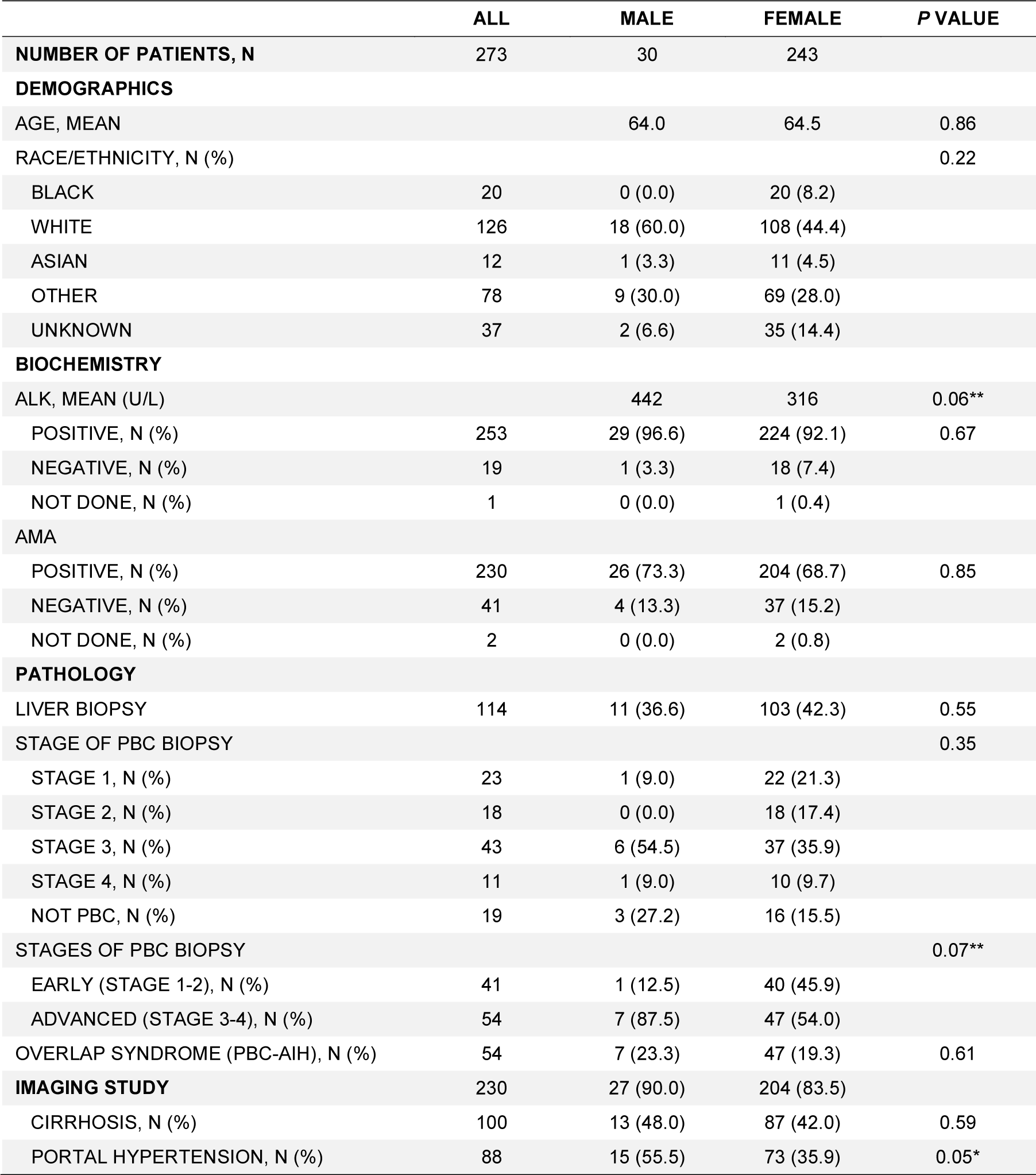
Comparison between Men and Women with PBC

### Comparison between AMA-positive and AMA-negative PBC

271 PBC patients were included for comparison. 230 (85%) had positive AMA and 41 (15%) had negative AMA (table 4). AMA-negative PBC patients had similar distributions of age, race/ethnicity and survival to AMA-positive PBC patients. 69 AMA-positive (30%) and 25 AMA-negative PBC patients (61%) had biopsy reports available in the Mount Sinai Health System. Biopsies done in outside systems were not included for analysis. The discrepancy in biopsy rates among AMA-positive and AMA-negative groups reflected the updated practice in making PBC diagnosis. Stage 1-2 was seen in 28/69 (41%) positive AMA vs. 13/25 (52%) negative AMA patients; stage 3-4 was observed in 41/69 (59%) positive AMA vs. 12/25 (48%) negative AMA patients. Imaging features were similar between AMA positive vs. negative PBC patients, including cirrhosis (91/208, 44% vs. 8/22, 36%), portal hypertension (64/208, 31% vs. 8/22, 36%) and abdominal lymphadenopathy (85/208, 41% vs. 5/22, 23%). Among 95 patients imaged with CT, lymphadenopathy was seen in 35/86 (41%) positive vs. 0/9 (0%) negative AMA patients (*p* < 0.05). However, this difference was not statistically significant in those imaged with MRI.

**Table 4.**
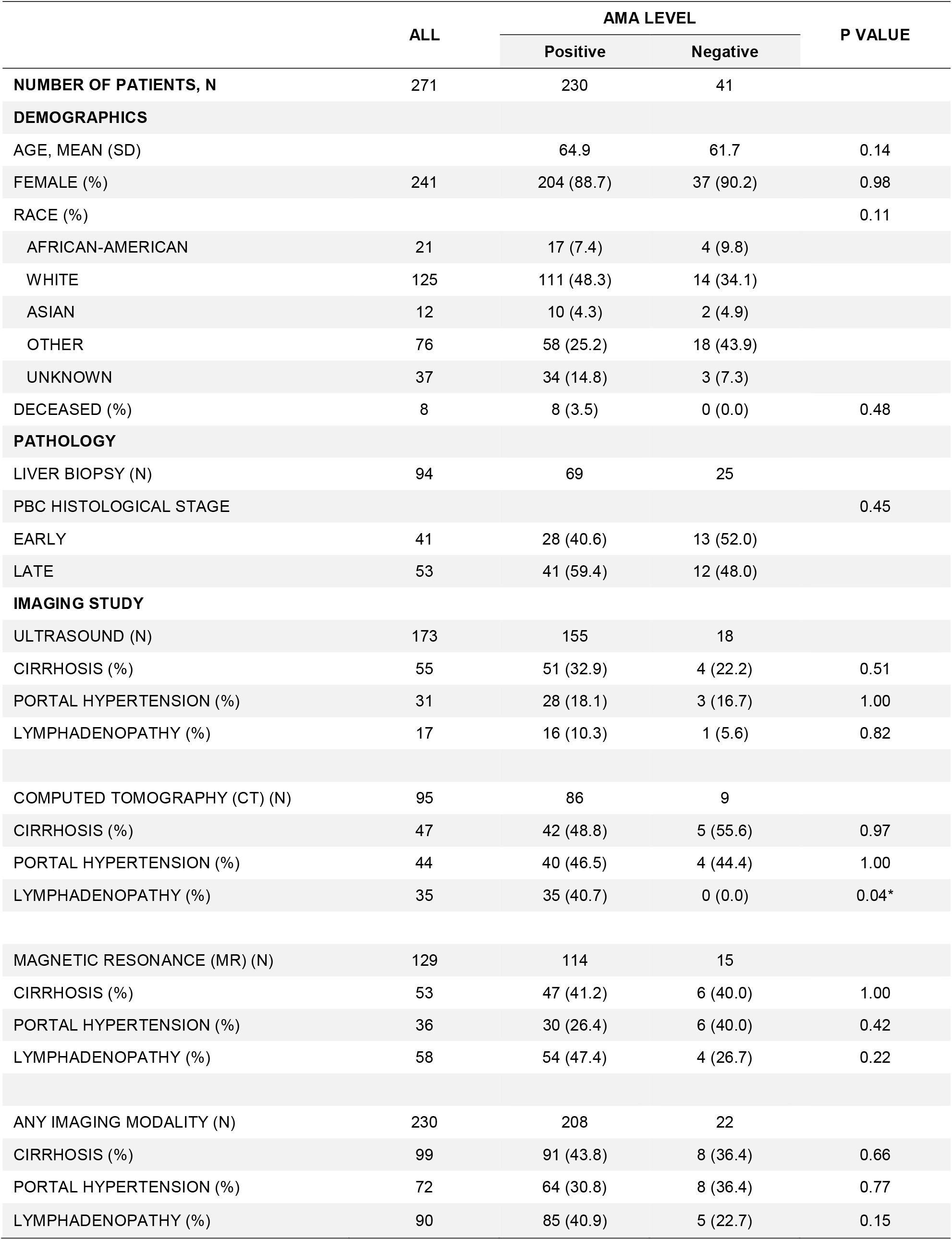
Comparison between AMA-positive and AMA-negative PBC.

|  | ALL | AMA LEVEL |  | P VALUE |
| --- | --- | --- | --- | --- |
|  |  | Positive | Negative |  |
| **NUMBER OF PATIENTS, N** | 271 | 230 | 41 |  |
| **DEMOGRAPHICS** |  |  |  |  |
| AGE, MEAN (SD) |  | 64.9 | 61.7 | 0.14 |
| FEMALE (%) | 241 | 204 (88.7) | 37 (90.2) | 0.98 |
| RACE (%) |  |  |  | 0.11 |
| AFRICAN-AMERICAN | 21 | 17 (7.4) | 4 (9.8) |  |
| WHITE | 125 | 111 (48.3) | 14 (34.1) |  |
| ASIAN | 12 | 10 (4.3) | 2 (4.9) |  |
| OTHER | 76 | 58 (25.2) | 18 (43.9) |  |
| UNKNOWN | 37 | 34 (14.8) | 3 (7.3) |  |
| DECEASED (%) | 8 | 8 (3.5) | 0 (0.0) | 0.48 |
| **PATHOLOGY** |  |  |  |  |
| LIVER BIOPSY (N) | 94 | 69 | 25 |  |
| PBC HISTOLOGICAL STAGE |  |  |  | 0.45 |
| EARLY | 41 | 28 (40.6) | 13 (52.0) |  |
| LATE | 53 | 41 (59.4) | 12 (48.0) |  |
| **IMAGING STUDY** |  |  |  |  |
| ULTRASOUND (N) | 173 | 155 | 18 |  |
| CIRRHOSIS (%) | 55 | 51 (32.9) | 4 (22.2) | 0.51 |
| PORTAL HYPERTENSION (%) | 31 | 28 (18.1) | 3 (16.7) | 1.00 |
| LYMPHADENOPATHY (%) | 17 | 16 (10.3) | 1 (5.6) | 0.82 |
| COMPUTED TOMOGRAPHY (CT) (N) | 95 | 86 | 9 |  |
| CIRRHOSIS (%) | 47 | 42 (48.8) | 5 (55.6) | 0.97 |
| PORTAL HYPERTENSION (%) | 44 | 40 (46.5) | 4 (44.4) | 1.00 |
| LYMPHADENOPATHY (%) | 35 | 35 (40.7) | 0 (0.0) | 0.04* |
| MAGNETIC RESONANCE (MR) (N) | 129 | 114 | 15 |  |
| CIRRHOSIS (%) | 53 | 47 (41.2) | 6 (40.0) | 1.00 |
| PORTAL HYPERTENSION (%) | 36 | 30 (26.4) | 6 (40.0) | 0.42 |
| LYMPHADENOPATHY (%) | 58 | 54 (47.4) | 4 (26.7) | 0.22 |
| ANY IMAGING MODALITY (N) | 230 | 208 | 22 |  |
| CIRRHOSIS (%) | 99 | 91 (43.8) | 8 (36.4) | 0.66 |
| PORTAL HYPERTENSION (%) | 72 | 64 (30.8) | 8 (36.4) | 0.77 |
| LYMPHADENOPATHY (%) | 90 | 85 (40.9) | 5 (22.7) | 0.15 |

## Discussion

PBC is a chronic, inflammatory, autoimmune disease of the biliary epithelial cells, causing slow progression of cholestasis and fibrosis^5^. The clinical and therapeutic aspects of PBC have been well studied, however relatively little is known about the radiographic features specific to PBC. Most previous studies have been conducted by radiologists with a focus on a single imaging modality and limited by small sample sizes. In addition, prior studies were based mainly on CT and MRI findings with close to none reporting typical US findings in PBC patients. To our knowledge this is the first large, single-center study outlining radiologic findings by ultrasound, CT and MRI, in PBC patients with early to advanced stages of disease.

Our study included 273 patients with PBC diagnosed per AASLD criteria, of which 89% were female, which is consistent with prior epidemiologic studies^2,12-15^. Out of the 273 patients, 172 underwent abdominal ultrasound, 95 underwent helical CT scan, and 129 underwent MRI/MRCP. We evaluated findings from these imaging modalities primarily because they are the most accessible and most commonly used in patients with liver pathology. US was able to detect most cases of cirrhosis and gallbladder abnormalities, while CT and MRCP were better at detecting lymphadenopathy, parenchymal pathology, biliary tree/pancreatic duct changes, and findings consistent with portal hypertension.

Findings consistent with cirrhosis were reported in 32%, 49.5%, and 41.1% of those who underwent abdominal US, CT, and MRI respectively. In one study looking at CT findings in 21 patients with PBC, cirrhosis was found in 43%, which is comparable to findings within our study^10^.

Recently, the periportal “halo sign” on T1- and T2-weighted MRI, characterized by rounded low signal-intensity abnormality around portal venous branches, has been identified as a specific finding in advanced PBC^3,11,16-18^. The prevalence of the halo sign has ranged from 27.6% to 75.8%, depending on the severity of disease in the cohorts analyzed. Our study reported a halo sign prevalence of 0.8%, a marked difference compared to prior findings. Our study retrospectively extracted imaging features from imaging reports, likely limiting observer bias on the part of the reporting radiologist and more likely reflecting the prevalence of this sign in “real world” practice.

We identified radiologic hepatomegaly in 9.3%, 8.4% and 10.1%, of patients who had abdominal US, CT, and MRI respectively. These findings are similar to what has been previously reported in other studies showing hepatomegaly in PBC patients with a prevalence ranging 11-19%^9,10^, and 37.8-56.5%^11,17,19^, for CT, and MRI respectively. Variations in hepatomegaly findings have been attributed to severity of disease with larger liver size associated with early disease.

Our cohort of patients did not have imaging findings consistent with HCC in any of the three imaging modalities. Indeterminate liver masses/cysts were identified in 8.7%, 17.9% and 35.7%, by US, CT, and MRI respectively. HCC in PBC is associated with more advanced fibrosis/cirrhosis^10^ and male sex^20^ - although the male sex association is likely a function of late diagnosis in men, rather than a more aggressive natural history of disease^14,21^. Our population had a smaller proportion of people with advanced PBC than previous studies reporting HCC in PBC, and this could explain the absence of HCC cases in our cohort.

Sequelae of portal hypertension were less prevalent in our cohort than previously reported^9,10,18,19^. Prior MRI studies have reported a prevalence of varices between 7.8-15.3%, splenomegaly between 20.1-86.4%, and ascites between 5.7-47.7%^11,16,22^. Portal hypertension is generally seen in more advanced disease and the wide range of distribution of these findings could be attributed to inherent differences in disease severity among the different patient cohorts. A CT-based study looking at sequelae of portal hypertension in pre-transplant patients (with more advanced/end-stage disease) reported 87%, 88%, and 44% prevalence of varices, splenomegaly and ascites respectively in their cohort^10^.

PBC results from autoimmune destruction of intrahepatic cholangiocytes leading to the obliteration of small bile ducts^3^. These biliary tree changes are more commonly reported on MRI/MRCP evaluations. In one study looking at MRI/MRCP findings in 13 patients with PBC, 8/13 (61.5%) had mild irregularity in the intrahepatic bile ducts and 1/13 (7.6%) was found to have focal narrowing at the common bile duct level^16^. This was comparable to the findings of biliary tree irregularity in 6.2% of MRI reports in our cohort. In another MRI-based study, 24/44 (54.5%) of PBC patients enrolled were found to have decreased number and stenosis of the intrahepatic bile ducts^22^. Our study found intrahepatic ductal dilatation in up to 7.4% and common bile duct dilatation (>7 mm) in up to 7.8% of patients, findings rarely reported previously in the literature. Like in our cohort, there is a paucity of US and CT studies in the literature reporting small biliary duct changes, likely a result of technical limitations with these modalities. We also reported unexpected features, such as cholelithiasis (up to 20%) and gallbladder wall thickening (up to 6.4%) within our cohort. Though these findings are non-specific for PBC, they may have some, as yet undescribed, significance in predicting severity and/or prognosis.

Lymphadenopathy can be a non-specific finding in infectious, inflammatory and malignant hepatic diseases^10,11,19^. Prominent or enlarged lymph nodes should always prompt a thorough malignancy workup, before being attributed to underlying PBC. The frequency and significance of lymphadenopathy in PBC remains controversial, but has been extensively reported in PBC radiology literature^7-9^. In our study, LAD was a frequent finding, noted in 11%, 58% and 69% of US, CT and MRI reports, respectively. This is comparable to the findings of several other studies, which report a wide range of frequencies of LAD^9,17,18,22^. In this study, lymphadenopathy was mostly reported in the periportal and portocaval regions and less frequently in the peripancreatic regions. The clinical significance of the presence, size or location of lymphadenopathy on imaging in PBC remains unclear, as there was no clear association between these variables and prognosis or stage of PBC in our study.

Men and women with PBC had similar clinical, histological and radiological features in our cohort, other than for the notable exception of a higher prevalence of portal hypertension in men. Though this may be related to a later diagnosis in men of this disease with a marked female preponderance^23^, we cannot exclude the possibility of a disease course that is more aggressive in men, or one in which portal hypertension develops earlier than would otherwise be expected. More work will be needed to investigate this possible link further, which, if confirmed, may make more frequent variceal screening in men advisable.

Our analysis found no evidence to suggest that AMA status (positivity or negativity) is associated with any distinct clinical, radiological or histological features. While it had been thought that AMA-negative PBC was characterized by a more aggressive course, findings from this large, tertiary center cohort do not support this hypothesis. Some other studies have also shown no difference in treatment response or overall clinical outcomes between AMA-negative and AMA-positive PBC patients^24-27^. It should be noted, however, that the rarity of AMA-negative PBC makes this a challenging entity to study, and there remains a risk of a type 2 statistical error in our work as a result of the relatively small pool of AMA-negative patients.

In terms of limitations, the retrospective nature of our study made it difficult to ensure that all data were available for all patients. For example, some patients had only one imaging modality available for inclusion, or no histology, or no direct temporal correlation between histology and radiology. This could bias findings by overrepresenting data from those who are more heavily investigated for important clinical, sociodemographic, or financial reasons. Secondly, it was not possible to accurately record the duration of follow-up and end outcome for each patient, which limited our ability to draw conclusions about the prognostic significance of some of our findings. For all imaging modalities, we depended on a single radiologist’s interpretation of each study and there was no second look verification of findings by another radiologist. This also limited the ability to expound on more nuanced radiologic findings such as extent of liver lobular atrophy/hypertrophy, caudate-to-right lobe ratio, and parenchymal changes that may be relevant in the evaluation and prognostication of PBC.

These limitations are outweighed by the benefit of a sample size larger than most prior studies, giving more insight into less common findings. Our study was conducted in a large health system in New York City, which includes five hospitals and a liver transplantation center. We included data from multiple radiology modalities instead of focusing on one modality as previous studies have done. We are unique in presenting ultrasound findings in PBC, which can guide clinicians diagnosing and managing PBC in settings where other radiologic modalities are unavailable or limited. Based on the distribution of PBC stage in the subset of patients who underwent biopsy, our study had a diverse distribution of patients across the whole spectrum of disease severity. Finally, our analysis of radiologic differences based on sex and AMA status are, to our knowledge, novel. Though there were no significant differences when we stratified by sex or AMA status, these findings are important and consistent with prior studies that note similar clinical presentation and prognosis in these subgroups.

## Data Availability

All data produced in the present study are available upon reasonable request to the authors

## Abbreviations

AASLD: American Association for the Study of Liver Diseases
AMA: Anti-mitochondrial antibodies
ALP: Serum alkaline phosphatase AIH Autoimmune hepatitis
CT: Computed tomography
EASL: European Association for the Study of the Liver
HCC: Hepatocellular carcinoma
IRB: Institutional Review Board
LAD: Lymphadenopathy
MRCP: Magnetic resonance cholangiopancreatography
MRI: Magnetic resonance imaging
PBC: Primary biliary cholangitis
PVH: Portal venus hypertension
SD: Standard deviation
US: Ultrasound

## Tables and figures

**Figure.**
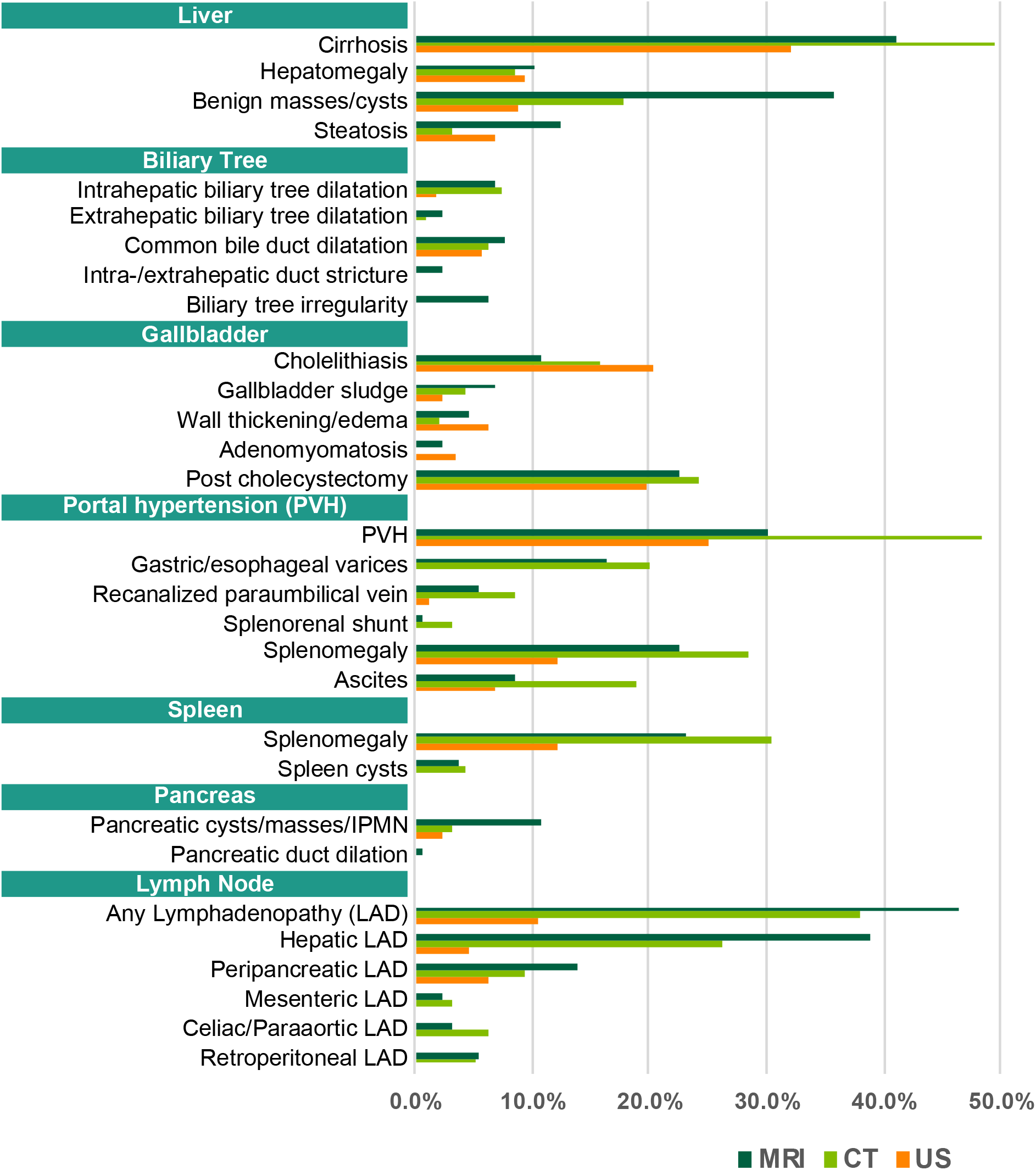
Imaging features of PBC patients.

## Reference

1. Kaplan MM, Gershwin ME. Primary biliary cirrhosis. N Engl J Med. Sep 22 2005;353(12):1261–73. doi:10.1056/NEJMra043898

2. Lleo A, Jepsen P, Morenghi E, et al. Evolving Trends in Female to Male Incidence and Male Mortality of Primary Biliary Cholangitis. Sci Rep. May 19 2016;6:25906. doi:10.1038/srep25906

3. Morgan MA, Sundaram KM. Primary biliary cholangitis: review for radiologists. Abdom Radiol (NY). Nov 7 2021;doi:10.1007/s00261-021-03335-x

4. European Association for the Study of the Liver. Electronic address eee, European Association for the Study of the L. EASL Clinical Practice Guidelines: The diagnosis and management of patients with primary biliary cholangitis. J Hepatol. Jul 2017;67(1):145–172. doi:10.1016/j.jhep.2017.03.022

5. Lindor KD, Bowlus CL, Boyer J, Levy C, Mayo M. Primary Biliary Cholangitis: 2018 Practice Guidance from the American Association for the Study of Liver Diseases. Hepatology. Jan 2019;69(1):394–419. doi:10.1002/hep.30145

6. Kobayashi S, Matsui O, Gabata T, et al. MRI findings of primary biliary cirrhosis: correlation with Scheuer histologic staging. Abdom Imaging. Jan-Feb 2005;30(1):71–6. doi:10.1007/s00261-004-0228-x

7. Dietrich CF, Leuschner MS, Zeuzem S, et al. Peri-hepatic lymphadenopathy in primary biliary cirrhosis reflects progression of the disease. Eur J Gastroenterol Hepatol. Jul 1999;11(7):747–53.

8. Eustace S, Buff B, Kane R, Jenkins R, Longmaid HE. The prevalence and clinical significance of lymphadenopathy in primary biliary cirrhosis. Clinical Radiology. 1995;50(6):396–399. doi:10.1016/s0009-9260(05)83137-x

9. Outwater E, Kaplan MM, Bankoff MS. Lymphadenopathy in primary biliary cirrhosis: CT observations. Radiology. Jun 1989;171(3):731–3. doi:10.1148/radiology.171.3.2717743

10. Blachar A, Federle MP, Brancatelli G. Primary biliary cirrhosis: clinical, pathologic, and helical CT findings in 53 patients. Radiology. Aug 2001;220(2):329–36. doi:10.1148/radiology.220.2.r01au36329

11. Idilman IS, Venkatesh SH, Eaton JE, et al. Magnetic resonance imaging features in 283 patients with primary biliary cholangitis. Eur Radiol. Sep 2020;30(9):5139–5148. doi:10.1007/s00330-020-06855-0

12. Flores A, Mayo MJ. Primary biliary cirrhosis in 2014. Curr Opin Gastroenterol. May 2014;30(3):245–52. doi:10.1097/MOG.0000000000000058

13. Galoosian A, Hanlon C, Zhang J, Holt EW, Yimam KK. Clinical Updates in Primary Biliary Cholangitis: Trends, Epidemiology, Diagnostics, and New Therapeutic Approaches. J Clin Transl Hepatol. Mar 28 2020;8(1):49–60. doi:10.14218/JCTH.2019.00049

14. John BV, Aitcheson G, Schwartz KB, et al. Male Sex Is Associated With Higher Rates of Liver-Related Mortality in Primary Biliary Cholangitis and Cirrhosis. Hepatology. Aug 2021;74(2):879–891. doi:10.1002/hep.31776

15. Podda M, Selmi C, Lleo A, Moroni L, Invernizzi P. The limitations and hidden gems of the epidemiology of primary biliary cirrhosis. J Autoimmun. Oct 2013;46:81–7. doi:10.1016/j.jaut.2013.06.015

16. Haliloglu N, Erden A, Erden I. Primary biliary cirrhosis: evaluation with T2-weighted MR imaging and MR cholangiopancreatography. Eur J Radiol. Mar 2009;69(3):523–7. doi:10.1016/j.ejrad.2007.11.003

17. Kovac JD, Jesic R, Stanisavljevic D, et al. Integrative role of MRI in the evaluation of primary biliary cirrhosis. Eur Radiol. Mar 2012;22(3):688–94. doi:10.1007/s00330-011-2296-y

18. Wenzel JS, Donohoe A, Ford KL, 3rd, Glastad K, Watkins D, Molmenti E. Primary biliary cirrhosis: MR imaging findings and description of MR imaging periportal halo sign. AJR Am J Roentgenol. Apr 2001;176(4):885–9. doi:10.2214/ajr.176.4.1760885

19. Meng Y, Liang Y, Liu M. The value of MRI in the diagnosis of primary biliary cirrhosis and assessment of liver fibrosis. PLoS One. 2015;10(3):e0120110. doi:10.1371/journal.pone.0120110

20. Abdulkarim M, Zenouzi R, Sebode M, et al. Sex differences in clinical presentation and prognosis in patients with primary biliary cholangitis. Scand J Gastroenterol. Nov 2019;54(11):1391–1396. doi:10.1080/00365521.2019.1683226

21. Rubel LR, Rabin L, Seeff LB, Licht H, Cuccherini BA. Does primary biliary cirrhosis in men differ from primary biliary cirrhosis in women? Hepatology. Jul-Aug 1984;4(4):671–7. doi:10.1002/hep.1840040418

22. Han D, He W. Associations Between Magnetic Resonance Imaging Findings and Scores of Liver Function and Histology in Patients with Primary Biliary Cirrhosis. Applied Magnetic Resonance. 2015;46(7):731–739. doi:10.1007/s00723-015-0675-2

23. Carbone M, Mells GF, Pells G, et al. Sex and age are determinants of the clinical phenotype of primary biliary cirrhosis and response to ursodeoxycholic acid. Gastroenterology. Mar 2013;144(3):560-569 e7; quiz e13-4. doi:10.1053/j.gastro.2012.12.005

24. Invernizzi P, Crosignani A, Battezzati PM, et al. Comparison of the clinical features and clinical course of antimitochondrial antibody-positive and -negative primary biliary cirrhosis. Hepatology. May 1997;25(5):1090–5. doi:10.1002/hep.510250507

25. Jin Q, Moritoki Y, Lleo A, et al. Comparative analysis of portal cell infiltrates in antimitochondrial autoantibody-positive versus antimitochondrial autoantibody-negative primary biliary cirrhosis. Hepatology. May 2012;55(5):1495–506. doi:10.1002/hep.25511

26. Juliusson G, Imam M, Bjornsson ES, Talwalkar JA, Lindor KD. Long-term outcomes in antimitochondrial antibody negative primary biliary cirrhosis. Scand J Gastroenterol. 2016;51(6):745–52. doi:10.3109/00365521.2015.1132337

27. Taylor SL, Dean PJ, Riely CA. Primary autoimmune cholangitis. An alternative to antimitochondrial antibody-negative primary biliary cirrhosis. Am J Surg Pathol. Jan 1994;18(1):91–9.

